# Identifying Racial Disparities in Clozapine Prescriptions Among Schizophrenia Patients using Data from Electronic Health Records

**DOI:** 10.1101/2022.11.17.22282446

**Authors:** Spenser Barry, L Fredrik Jarskog, Kai Xia, Rohit Simha Torpunuri, Xiaoyu Wu, Xiaoming Zeng

## Abstract

**Objective:** This study sought to assess the demographic factors that alter the likelihood of being prescribed clozapine. The primary hypothesis is that African American patients are less likely to be prescribed Clozapine than White and other racial groups. Additionally, this study aims to expand on earlier studies of clozapine by examining how multiple variables, especially social determinants of health, gender, rurality, and care patterns affect the rates at which clozapine is prescribed.

**Methods:** This observational study examines the racial disparities and other factors associated with receiving a clozapine prescription for patients with a schizophrenia diagnosis using structured data in the Electronic Health Records (EHR) from a multi-facility health system from 10/1/2015 - 11/30/2021. Bivariate analysis, multiple logistic regression, and sensitivity analysis tests were performed to determine which factors differed significantly between the group of patients who were prescribed clozapine and those who were not.

**Results:** Results showed that being white and having a higher socioeconomic income was associated with an increased clozapine prescription rate, while gender, rurality, age at first diagnosis, and ethnicity did not influence prescription likelihood. Increased treatment length was also associated with a greater likelihood of being prescribed clozapine.

**Conclusion:** African American patients are over-diagnosed with schizophrenia and under-prescribed clozapine compared to Caucasians after controlling for the variables associated with demographics, social determinants of health, and care access patterns. Future research is needed to understand and disentangle the biological, clinical, administrative, and societal causes behind the disparity in clozapine treatment.

## Introduction

Schizophrenia affects approximately 1.5 million people in the United States [1], and about 30% of these patients are estimated to have Treatment-resistant schizophrenia (TRS) [2]. Clozapine is considered the gold standard for TRS; however, it remains underutilized in clinical settings in the U.S [3].

Previous literature has suggested that several demographic factors may influence clozapine prescription rates. It is well documented that being African American reduces patients’ odds of being prescribed clozapine [4-8]. These studies reported a wide variety of odds ratios comparing clozapine prescription rates from a white patient to a black patient—ranging from 1.35 to 6.00, which indicates that a white patient will have a higher chance of being prescribed clozapine compared to a black patient.

It remains unclear why clozapine prescriptions vary by race and why minorities are under-prescribed clozapine. Examining biological factors indicates that black patients have a lower white blood cell count and a higher risk of developing neutropenia than their white counterparts. This may result in more treatment interruptions and higher discontinuation of clozapine [9]. Aside from biological differences, several other reasons for the disparity in prescription rates have been suggested, such as lack of prescriber experience, patient distrust of clinicians, and systematic barriers in medicine; however, the effect of these factors remains uncertain and needs further exploration [10].

Previous literature has shown mixed evidence that gender influences clozapine prescription. A retrospective study of 2244 patients from South London examined patients with treatment-resistant schizophrenia. It showed that men were prescribed clozapine in 85% of cases, compared to women’s 77%, representing a statistically significant difference between the groups [11]. Furthermore, a retrospective study in the United States of 326,119 patients, using Medicaid Claims data, found that compared to women, men had an odds ratio of 1.26 for clozapine prescriptions [12]. However, other studies have provided evidence that gender does not influence relative clozapine prescription, Martin and colleagues examined how socioeconomic status influences a patient’s odds of being prescribed clozapine. They studied 3,200 patients in Glasgow, Scotland, examining if socioeconomic status, calculated by approximating patient wealth based on postcodes, predicted the rates at which patients were prescribed depot antipsychotic medication and antipsychotics, including clozapine [13]. Their study found no relationship between a patient’s socioeconomic status and their likelihood of being prescribed clozapine.

Our study will add to the existing literature by providing further proof of a racial treatment disparity between black and white schizophrenia patients by using data from an Electronic Health Record (EHR) system. The additional novelty includes showing that Social Determinants of Health (SDOH) will have predictive validity for determining clozapine prescription likelihood, which, to our knowledge, no other study has demonstrated. Furthermore, we provide evidence against the influence of gender, rurality, ethnicity, and age at first diagnosis for impacting clozapine prescriptions. This research is vital to improve the understanding of the factors influencing clozapine prescription and inform clinicians and researchers about the treatment disparities between racial groups.

## RESEARCH QUESTIONS

The primary hypothesis of the study is that African American patients are less likely to be prescribed Clozapine than White and other racial groups. Additionally, this study aims to expand on earlier studies of clozapine by examining how multiple variables, especially other demographic factors, SDOH, and care patterns affect the rates at which clozapine is prescribed.

## METHODS

### Patient Selection

This retrospective cross-sectional study used data from the University of North Carolina Health’s Electronic Health Records system. UNC Health is the academic health system for the School of Medicine at the University of North Carolina-Chapel Hill. We extracted structured encounter and prescription data of 3159 adult schizophrenia patients (age:18-64, primary diagnosis ICD-10 codes: F20) between 10/1/2015 and 11/30/2021. No patient deaths were reported at the time of our data extraction. The study was reviewed and approved by the UNC-CH Institutional Review Board (IRB # 21-1654).

### Variables

### Outcome variable

The outcome variable of the study is whether a Schizophrenia patient had been prescribed clozapine during the study period. Prescription duration and dosage data were not factored in the coding of the clozapine prescription.

### Predictor variable

The patient’s race is the primary predictor variable in the study, coded as Black, White, and Others.

### Covariables

#### Demographic variables

1. Gender: Female vs. male.
2. Ethnicity: Hispanic or Latino, not Hispanic or Latino, Unspecified
3. Age at the earliest recorded diagnosis
4. Rurality: determined by matching the patient’s Census Tract with the USDA’s 2010 Revised Rural-Urban Commuting Area Codes (RUCA) [14]. A census tract with a RUCA score > 4 is deemed rural, otherwise urban.

#### Social Determinants of Health Variables

The extent of SDOH for each patient was determined using the 2018 census tract level Social Vulnerability Index (SVI). SVI is a tool developed by the Centers for Disease Control and Prevention (CDC) to measure the extent of social vulnerability within a county or census tract[15]. An increasing number of studies have employed the SVI index as a proxy for SDOH [16, 17]. The SVI score is a composite measure combining and normalizing the scores of four different themes derived from percentile ranking U.S. census data of different variables at the corresponding geographic level – census tract or county. Higher SVI scores correlate with more vulnerability for the patients. The four thematical SVI scores are also included in the analysis -- SVI Theme 1 (Socioeconomic Status), SVI Theme 2 (Household Composition and Disability), SVI Theme 3 (Minority Status and Language), SVI Theme 4 (Housing and Transportation).

#### Care Pattern Variables

1. Number of Years in the Treatment – the number of years between the first and last encounters for patients with a primary diagnosis of schizophrenia.
2. Number of Different Non-Clozapine Antipsychotics – the number of different classes of antipsychotics, other than clozapine, that was prescribed during the study period for each patient.
3. Average Number of Encounters per year – The total number of encounters divided by the number of years in the treatment.

### Data Analysis

Descriptive statistics were performed on all variables of interest. We used a two-sample t-test for continuous variables and *χ*^2^ test for categorical variables for the bivariate analysis.

A multiple logistic regression analysis was conducted to examine the association between clozapine prescription and the patient’s race, controlling for the other variables in the model. We removed the overall SVI score variable in the logistic regression model to minimize collinearity.

Because 303 patients have missing values on antipsychotic treatment from the extracted data and were not included in the primary analysis, we conducted a sensitivity analysis by replacing the missing values with zero for these patients. The possible reasons for the missing data vary: some patients may have decided on alternative treatment other than medication for their conditions or sought care elsewhere. The literature has reported that 70% of patients with schizophrenia spectrum disorders in remission might not engage in active pharmacotherapy [18].

Data were pre-processed using Tableau Prep (version 2022.11.1) and analyzed the data using R (version 4.2.1). Unless specified otherwise, alpha is set at 0.05 for statistical significance in the analysis.

## RESULTS

### Descriptive Statistics

Table 1 shows the univariate and bivariate statistics of the variables. In the dataset of 3159 schizophrenia patients, 2855 had antipsychotics prescription data. Of those patients, 401 (14%) received a prescription for Clozapine during the study period. Notably, the percentage of patients prescribed clozapine is lower than the 30% estimated prevalence of TRS among schizophrenia patients.

**Table 1.**
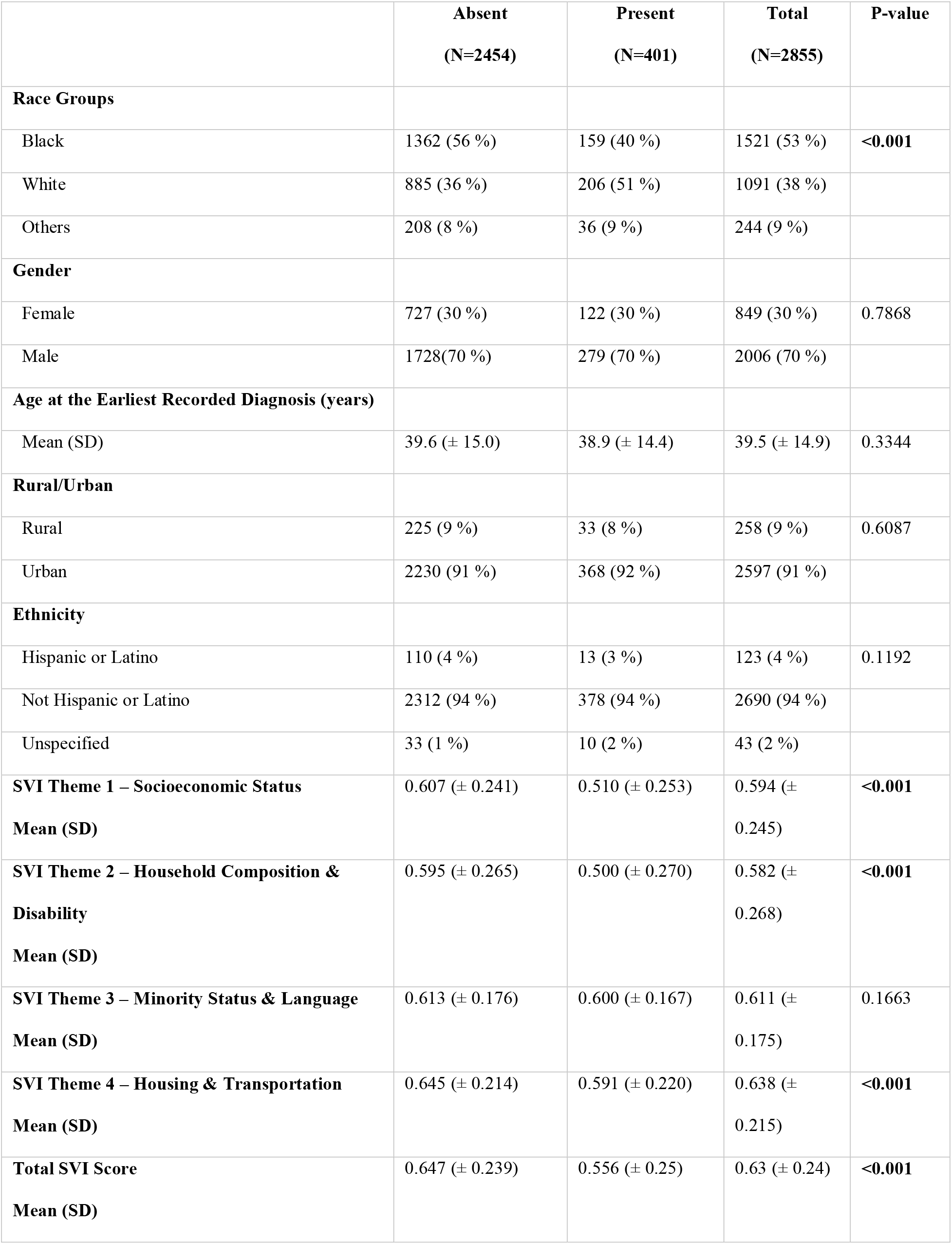
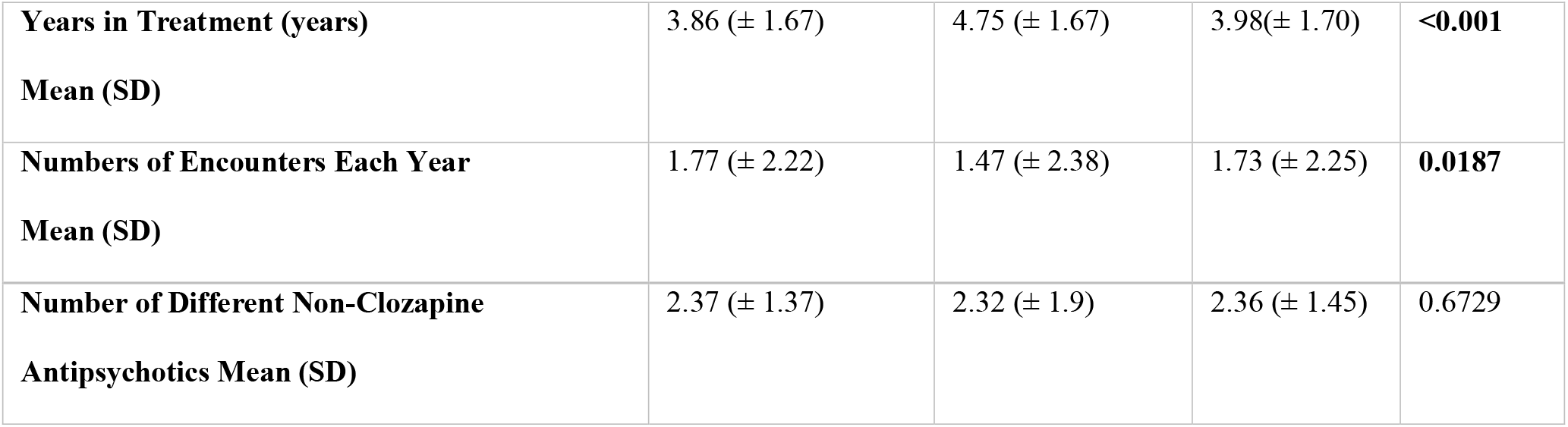
Characteristics of Patients who were prescribed or not prescribed clozapine.

70% of the patients were male, and the average age of the first diagnosis recorded in our dataset was 39.5 (SD 14.9). Most (91%) of the patients live in an urban setting. 94% of the population was not Hispanic or Latino. The four thematic SVI scores and the overall SVI score all fell within the moderate-high range (0.5-0.75) quartile.

The average number of years in treatment was 3.98 years (SD 1.70). On average, a patient had 1.73 visits (SD 2.25) per year to UNC for their schizophrenia. Excluding clozapine, patients received an average of 2.36 (SD 1.45) other types of antipsychotics during the study period.

### Bivariate Analysis

Among the schizophrenia patients, 1521 (53%) were black, and 1090 (38%) were white – a 15% difference. Like previous studies, our study showed that African Americans make up a much larger percentage of the schizophrenic population compared to the general racial breakdown of the population in North Carolina [19]. However, among the 401 patients who received clozapine prescriptions, 53% were white and 40% were black, which is the opposite of the racial pattern shown in the diagnosis of schizophrenia.

The binary analysis showed that 10.4% (159/1521) of black patients had been prescribed clozapine, compared to the prescription rate of 18.8% (206/1091) for white patients during the same study period. χ^2^ test results showed a statistically significant difference between racial groups (p<0.001). No other demographic variables – gender, age at the first recorded diagnosis, rurality, or ethnicity are related to clozapine prescription – had statistical significance.

The overall SVI score of the clozapine patients was 9.1 percentiles lower than that of non-clozapine patients, with a two sample t-test showing statistically significant differences (p<0.001). Among the four SVI themes, non-clozapine patients had lower scores in all areas compared to the clozapine patients, with three themes (themes 1, 2, and 4) showing statistically significant differences.

Among the care access pattern variables in the bivariate analysis, both variables *Years in Treatment* and the *Number of Encounters Each Year* were statistically significant between the clozapine present and absent groups. On average, the clozapine present group was treated for a longer period of time than the non-clozapine group (4.75 vs. 3.86 years) but visited the providers less often (1.47 vs. 1.77 visits per year). No statistical significance was found between the two groups for the variable *Number of Different Non-Clozapine Antipsychotics*.

## Multiple Logistic Regression

Table 2 shows the results of multiple logistic regression. Compared to black patients, white patients are 71.3% (OR, 1.713) more likely to be prescribed clozapine after controlling for other variables. No other demographic variables showed statistical significance, consistent with the bivariate analysis. SVI themes 1 and 3 variables are statistically significant in the logistic regression model. *Years in Treatment* remains a statistically significant variable (OR = 1.355) for clozapine prescription. No other care pattern variables were statistically significant.

**Table 2.** Odds Ratios of the Logistic Regression Model

| Characteristic | OR<sup>1</sup> | 95% CI<sup>1</sup> | p-value |
| --- | --- | --- | --- |
| **Racial Groups** |  |  | **<0.001** |
| Black | — | — |  |
| White | 1.713 | 1.343, 2.187 |  |
| Others | 1.397 | 0.866, 2.204 |  |
| **Rural/Urban** |  |  | 0.4 |
| Rural | — | — |  |
| Urban | 0.843 | 0.566, 1.287 |  |
| **Gender** |  |  | 0.6 |
| Female | — | — |  |
| Male | 0.930 | 0.730, 1.191 |  |
| **Ethnicity** |  |  | 0.11 |
| Hispanic or Latino | — | — |  |
| Not Hispanic or Latino | 1.493 | 0.791, 3.007 |  |
| Unspecified | 2.772 | 1.062, 7.104 |  |
| **SVI Theme 1 Score** | 0.271 | 0.122, 0.597 | **0.001** |
| **SVI Theme 2 Score** | 0.85 | 0.473, 1.541 | 0.6 |
| **SVI Theme 3 Score** | 2.972 | 1.336, 6.720 | **0.007** |
| **SVI Theme 4 Score** | 0.862 | 0.441, 1.689 | 0.7 |
| **No. of Different Non-clozapine Antipsychotics** | 0.97 | 0.900, 1.047 | 0.5 |
| **Numbers of Encounters Each Year** | 1.055 | 0.996, 1.108 | 0.066 |
| **Age at the Earliest Recorded Diagnosis** | 0.997 | 0.99, 1.004 | 0.4 |
| **Years in Treatment** | 1.355 | 1.258, 1.460 | **<0.001** |
| <sup>1</sup> OR = Odds Ratio, CI = Confidence Interval |  |  |  |

In the sensitivity analysis, the primary input variable – racial group, remains statistically significant in both the bivariate and logistic regression analyses. In the binary analysis, the variable “number of different non-clozapine antipsychotics” became statistically significant between the clozapine present and clozapine absent groups. This statistical significance may have reversed because the effect of the imputation is to treat the patients with missing antipsychotic values as not receiving any antipsychotic treatment, including clozapine, which consequently increased the sample size for the non-clozapine group. The variable “Number of Encounters Each Year” became statistically insignificant in the sensitivity analysis. However, the p-value (0.0508) from the t-test is just slightly above the alpha level. On average, the number of annual encounters in the clozapine absent group decreased by 0.1, which again is the effect of replacing missing values with a zero imputation in the data.

In the multiple logistic regression, the p-value of the variable *Number of Encounters Each Year* changed from 0.045 to 0.051; however, it remains close to the alpha value (0.05). No statistical significance was changed for other variables. Overall, we are confident that our initial results are robust enough after replacing missing values with zeros for patients who miss the antipsychotic information and comparing results.

Details of the sensitivity analysis are included in the supplemental document.

## DISCUSSION

In this cross-sectional observational study, we used the 6-year (2015-2021) real-world data from the EHR of a large healthcare system to assess the racial disparities in clozapine prescriptions by comparing the African American group with White and other racial groups. Our primary finding is that racial disparities exist in prescribing clozapine to schizophrenia patients, even after controlling for other demographic, SDOH, and care pattern variables. Being African American and having a higher SVI score decreased the likelihood of a patient being prescribed clozapine. The reasons for the under-prescription of clozapine in black schizophrenia patients could be multiple–prescriber bias, the anticipation of nonadherence, and the notion of lower effectiveness in African Americans [20].

One novel approach to our study is incorporating the SVI – a measurement of SDOH in our analysis. It is worth highlighting that all the patients’ thematic and overall SVI scores fall in the moderate to high (0.5-0.75) quartile, reflecting the general social vulnerability of the patient cohort in the study. Both the overall SVI score and the four thematic SVI scores are higher in the non-clozapine antipsychotic group than in the clozapine group, which was expected because patients need more resources to access care to maintain long-term clozapine treatment.

Among all schizophrenia patients in the entire UNC Health System, from 2015 to 2021, the prescription rate for clozapine was about 14%. It is much higher than previously estimated in the state of North Carolina [21]. One organizational fact is that the Center for Excellence in Community Mental Health (CECMH) at UNC Health provides evidence-based care to patients with Serious Mental Illness (SMI). The internal dashboard reported that the clozapine prescription rate at CECMH is 16.4% [22], even higher than the UNC average. This demonstrates the value of a specialized SMI community mental health center in promoting and implementing evidence-based psychiatric care in academic and community settings [23].

No statistically significant differences were found between the status of clozapine prescription and patients’ gender, age, ethnicity, and rural/urban status. From the clinical visit pattern variables, only the years in treatment significantly impact clozapine prescription. This should not be a surprise – the longer a patient is under treatment, the more likely the patient will develop resistance to non-clozapine antipsychotics and, consequently, will be prescribed clozapine.

We also found that African Americans are the majority racial group diagnosed with schizophrenia, 53% vs. 38% white. Compared to the general racial breakdown of North Carolina, where an estimated 22% of residents are black and 70% are white, these results suggest that black patients are much more likely to be diagnosed with schizophrenia than their white counterparts [24]. The finding matches results from existing literature. For example, a meta-analysis published in 2018 found that African American patients are 2.4 times more likely to be diagnosed with schizophrenia [25]. Their study suggested that the disproportionally higher diagnosis of schizophrenia patients might be caused by multiple reasons, including genetic, environmental, socioeconomic, and clinical reasoning [26].

The combination of overdiagnosis of schizophrenia in the black population and under-prescription of clozapine among black schizophrenia patients results in a gap in care quality for African American patients. According to past epidemiological studies of Medicaid schizophrenia patients, clozapine prescription rates have been comparatively lower in North Carolina than the U.S. average [27]. Early alerting to clinicians will be critical to addressing this patient group’s subpar prescription of clozapine.

Our study found that, on average, patients received 2.36 different types of antipsychotics besides clozapine, which is over the 2 non-clozapine antipsychotic trials indicated in the FDA’s guidelines before clozapine should be prescribed [28]. However, since our study is cross-sectional, we could not establish the temporal relationship between the prescriptions of non-clozapine antipsychotics and clozapine or estimate the number of non-clozapine prescriptions before clozapine. Future studies will be needed to build a sequential relationship between clozapine and other types of antipsychotics.

Our study found no gender difference in clozapine prescription rates. As previously discussed, this finding is consistent with some studies but not with others. However, our study did find a gender disparity in the diagnosis of schizophrenia consistent with other literature. In 1992, Nicole and colleagues reported an overall female: male schizophrenia diagnosis ratio of 1: 1.77. Our study found a higher female: male diagnosis ratio than their study, reporting a ratio of 1: 2.33 (30% female vs. 70% male). Compared to females, the percentage of males diagnosed with schizophrenia in our dataset is significantly higher than gender breakdown of North Carolina [30]. Additional data may be needed to confirm the finding fully.

It surprised us that there is no difference between the rural and urban populations regarding clozapine prescription. However, given that more minorities live in urban areas, the significant difference between the racial groups may overpower the difference between urban and rural groups.

### Limitations

Due to its cross-sectional and single-site design, the study’s outcome may not be generalizable to other geographic areas. However, given the statewide coverage of the UNC Health system, we are confident that our data reflects the racial disparities in the state of North Carolina. Future studies should leverage the existing networks of observational data platforms to increase generalizability.

Due to the scope of the data extraction, we did not obtain data on other patient-level variables, notably other mental health disorders such as substance abuse or diabetes. Future studies need to address these limitations by expanding the data query to include comorbidities.

We used the data from EHR as the basis for the analysis, which has gained momentum in clinical research due to the wide adoption of EHR systems and access to “big data” [31, 32]. However, inheritably such data has limitations that need to be adequately considered in mental health service research.

Census tract level SVI scores were used to quantify the population’s social determinants of health. As SVI scores are a measure of population health, any individual patient could be distinct from the average residents in the same census tract, causing an imprecise estimate. Further research could improve our study’s results by using patient-level data to examine SDOH.

## CONCLUSIONS

Our study found that racial disparities exist in clozapine prescription and schizophrenia diagnosis using data from an Electronic Health Record System from 2015-2021. Analysis showed that African American patients are over-diagnosed with schizophrenia and under-prescribed clozapine compared to Caucasians after controlling for the variables associated with demographics, SDOH, and care access patterns. Future research needs to explore and disentangle the biological, clinical, administrative, and societal causes behind the disparity in clozapine treatment.

### Disclosures

This work was supported by a 2021 grant from the foundation of hope. The authors report no conflicting obligations at this time.

## Data Availability

All data produced in the present study are available upon reasonable request to the authors

