## Supplement Table 1 & 2 for "Identifying Racial Disparities in Clozapine Prescriptions Among Schizophrenia Patients using Data from Electronic Health Records"

Table 1: Descriptive statistics of variables

| | **Absent (N=2759)** | **Present (N=401)** | **Total (N=3160)** | **P-value** |
|---|---|---|---|---|
| Race_Groups | | | | |
| Black | 1497 (54 %) | 159 (40 %) | 1656 (52 %) | < 0.001 |
| White | 1026 (37 %) | 206 (51 %) | 1232 (39 %) | |
| Others | 234 (8 %) | 36 (9 %) | 270 (9 %) | |
| Gender | | | | |
| Female | 837 (30 %) | 122 (30 %) | 959 (30 %) | 1 |
| Male | 1922 (70 %) | 279 (70 %) | 2201 (70 %) | |
| Age at the Earlist Recorded Diagnosis (years) | | | | |
| Mean (SD) | 40.4 (± 15.3) | 38.9 (± 14.4) | 40.2 (± 15.2) | 0.05156 |
| Rural/Urban | | | | |
| Rural | 269 (10 %) | 33 (8 %) | 302 (10 %) | 0.3783 |
| Urban | 2488 (90 %) | 368 (92 %) | 2856 (90 %) | |
| Ethnicity | | | | |
| Hispanic or Latino | 115 (4 %) | 13 (3 %) | 128 (4 %) | 0.3764 |
| Not Hispanic or Latino | 2595 (94 %) | 378 (94 %) | 2973 (94 %) | |
| Unspecified | 47 (2 %) | 10 (2 %) | 57 (2 %) | |
| SVI Theme 1 Score | | | | |
| Mean (SD) | 0.610 (± 0.240) | 0.510 (± 0.253) | 0.597 (± 0.244) | < 0.001 |
| SVI Theme 2 Score | | | | |
| Mean (SD) | 0.603 (± 0.265) | 0.500 (± 0.270) | 0.590 (± 0.268) | < 0.001 |
| SVI Theme 3 Score | | | | |
| Mean (SD) | 0.606 (± 0.179) | 0.600 (± 0.167) | 0.605 (± 0.178) | 0.5314 |
| SVI Theme 4 Score | | | | |
| Mean (SD) | 0.646 (± 0.214) | 0.591 (± 0.220) | 0.639 (± 0.216) | < 0.001 |
| Total SVI Score | | | | |
| Mean (SD) | 0.649 (± 0.238) | 0.556 (± 0.254) | 0.638 (± 0.242) | < 0.001 |
| Years in Treatment (years) | | | | |
| Mean (SD) | 3.85 (± 1.67) | 4.75 (± 1.67) | 3.97 (± 1.70) | < 0.001 |
| Numbers of Encounters Each Year | | | | |
| Mean (SD) | 1.72 (± 2.14) | 1.47 (± 2.38) | 1.68 (± 2.17) | 0.05156 |
| No. of Different Non-clozapine Antipsychotics | | | | |
| Mean (SD) | 2.11 (± 1.49) | 2.32 (± 1.87) | 2.13 (± 1.55) | 0.02565 |

Table 2: Results of Multiple Logistic Regression

| **Characteristic** | **OR[1]** | **95% CI[1]** | **p-value** |
| --- | --- | --- | --- |
| Race_Groups | | | **<0.001** |
| Black | — | — | |
| White | 1.680 | 1.320, 2.141 | |
| Others | 1.314 | 0.820, 2.060 | |
| Rural/Urban | | | 0.4 |
| Rural | — | — | |
| Urban | 0.846 | 0.572, 1.286 | |
| Gender | | | 0.7 |
| Female | — | — | |
| Male | 0.96 | 0.755, 1.225 | |
| Ethnicity | | | 0.2 |
| Hispanic or Latino | — | — | |
| Not Hispanic or Latino | 1.428 | 0.762, 2.859 | |
| Unspecified | 2.410 | 0.938, 6.061 | |
| SVI Theme 1 Score | 0.256 | 0.118, 0.556 | **<0.001** |
| SVI Theme 2 Score | 0.704 | 0.398, 1.254 | 0.2 |
| SVI Theme 3 Score | 3.983 | 1.810, 8.919 | **<0.001** |
| SVI Theme 4 Score | 0.861 | 0.444, 1.673 | 0.7 |
| No. of Different Non-clozapine Antipsychotics | 1.069 | 0.997, 1.144 | 0.060 |
| Numbers of Encounters Each Year | 1.053 | 0.99, 1.106 | 0.073 |
| Age at the Earlist Recorded Diagnosis | 0.99 | 0.99, 1.003 | 0.2 |
| Years in Treatment | 1.345 | 1.250, 1.448 | **<0.001** |

[1] OR = Odds Ratio, CI = Confidence Interval
